# Assessment of cardiovascular & pulmonary pathobiology *in vivo* during acute COVID-19

**DOI:** 10.1101/2022.03.21.22272698

**Authors:** Shirjel R Alam, Sudhir Vinayak, Adeel Shah, Gemina Doolub, Redemptar Kimeu, Kevin P Horn, Stephen R. Bowen, Mohamed Jeilan, Ken Lee, Sylvia Gachoka, Felix Riunga, Rodney D. Adam, Hubert Vesselle, Nikhil Joshi, Mariah Obino, Khalid Makhdomi, Kevin Onyinkwa, Edward Nganga, Samuel Gitau, Michael Chung, Anoop S V Shah

## Abstract

**Importance:** Acute COVID-19-related myocardial, pulmonary and vascular pathology, and how these relate to each other, remains unclear. No studies have used complementary imaging techniques, including molecular imaging, to elucidate this.

**Objective:** We used multimodality imaging and biochemical sampling *in vivo* to identify the pathobiology of acute COVID-19.

**Design, Setting and Participants:** Consecutive patients presenting with acute COVID-19 were recruited during hospital admission in a prospective cross-sectional study. Imaging involved computed-tomography coronary-angiography (CTCA - identified coronary disease), cardiac 2-deoxy-2-[fluorine-18]fluoro-D-glucose positron-emission tomography/computed-tomography (18F-FDG-PET/CT - identified vascular, cardiac and pulmonary inflammatory cell infiltration) and cardiac magnetic-resonance (CMR – identified myocardial disease), alongside biomarker sampling.

**Results:** Of 33 patients (median age 51 years, 94% male), 24 (73%) had respiratory symptoms, with the remainder having non-specific viral symptoms. Nine patients (35%, n=9/25) had CMR defined myocarditis. 53% (n=5/8) of these patients had myocardial inflammatory cell infiltration. Two patients (5%) had elevated troponin levels. Cardiac troponin concentrations were not significantly higher in patients with myocarditis (8.4ng/L [IQR 4.0-55.3] vs 3.5ng/L [2.5-5.5], p=0.07) or myocardial cell infiltration (4.4ng/L [3.4-8.3] vs 3.5ng/L [2.8-7.2], p=0.89). No patients had obstructive coronary artery disease or vasculitis. Pulmonary inflammation and consolidation (percentage of total lung volume) was 17% (IQR 5-31%) and 11% (7-18%) respectively. Neither were associated with presence of myocarditis.

**Conclusions and relevance:** Myocarditis was present in a third patients with acute COVID-19, and the majority had inflammatory cell infiltration. Pneumonitis was ubiquitous, but this inflammation was not associated with myocarditis. The mechanism of cardiac pathology is non-ischaemic, and not due to a vasculitic process.

**Key Points:** *Question:* What is the pathobiology of the cardiac, pulmonary and vascular systems during acute COVID-19 infection ?

*Findings:* Over a third of patients with acute COVID-19 had myocarditis by cardiac MRI criteria. Myocardial inflammatory cell infiltration was present in about two thirds of patients with myocarditis. No associations were observed between the degree of pulmonary involvement and presence of myocarditis. There was no evidence of obstructive coronary artery disease or evidence of large vessel vasculitis.

*Meaning:* Myocarditis is common in acute COVID-19 infection, and may be present in the absence of significant pulmonary involvement. The cause of myocarditis is inflammatory cell infiltration in the majority of cases, but in about a third of cases this is not present. The mechanism of cardiac pathology in acute COVID-19 is non-ischaemic, and vascular thrombosis in acute COVID-19 is not due to significant coronary artery disease or a vasculitic process.

## INTRODUCTION

The coronavirus disease 2019 (COVID-19) has been mostly associated with pulmonary injury, but its association with cardiac and vascular pathobiology remains poorly understood (1-3). Patients with cardiac involvement are at a higher risk of mortality, with 8-28% of patients showing biochemical evidence of myocardial injury (4).

2-deoxy-2-[fluorine-18]fluoro-D-glucose positron emission tomography integrated with computed tomography (18F-FDG-PET/CT) can identify cellular inflammation in pulmonary, cardiac and vascular tissues, but prospective studies in COVID-19 remain limited (5-7). Whilst cardiac magnetic resonance (CMR) (1-3) and chest computed tomography (CT) imaging in COVID-19 have been conducted (8) these have been limited to the recovery phase and restricted to a single modality. As such these studies were unable to differentiate ischaemic from non-ischaemic cardiac pathology. A multisystem inflammatory syndrome in children (MIS-C) with myocarditis and cardiac impairment as hallmarks of the presentation has been described (9). Whether a similar mechanism of cardiac and vascular injury occurs in the adults with acute COVID-19 remains unknown.

Using CMR, computed tomography coronary angiography (CTCA) (10) and 18F-FDG-PET/CT (5-7) imaging during *acute* COVID-19 infection, we investigated *in vivo* pathobiology of the myocardium, arterial vasculature and pulmonary parenchyma. We hypothesised that myocardial or pulmomary inflammation and injury could be described by CMR and 18F-FDG-PET/CT, the presence of vascular inflammation identified by 18F-FDG-PET/CT and the contribution by coronary artery disease shown by CTCA. We investigated the relationship between imagining findings and biomarkers, as well as any association between pulmonary and cardiac pathology.

## METHODS

### Study design and population

Participants hospitalised with COVID-19 at the Aga Khan University Hospital in Nairobi, Kenya were recruited in this single-centre exploratory observational study. The full study methodology with the inclusion and exclusion criteria and imaging techniques and protocols has been pubsished (11). The study complies with the Declaration of Helsinki with study approval from the Aga Khan University Nairobi Institutional Ethics Review Committee (Reference: 2020/IERC-74 (v2).

Excluision criteria were contra-indication to CMR, known previous myocardial pathology, and those with severe symptoms requiring non-invasive or invasive ventilation. Patients underwent multi-modality imaging and serological testing for High-sensitivity cardiac troponin-I (hsTrop, Siemens Healthineers), N-terminal pro B-type natriuretic peptide (NT-proBNP, Siemens Atellica Solution), C-Reactive protein (CRP, Siemens Atellica Solution) and viral load (12) (using cycle threshold [CTVL], RealStar® SARS-CoV-2 RT-PCR Kit, Altona Diagnostics. We additionally identified a small prospective control population of individuals where COVID-19 was excluded, and age- and sex-matched historical control population who had previously undergone 18F-FDG-PET/CT.)

### Image Acquisition & Assessment

Participants underwent simultaneous CTCA and thoracic 18F-FDG-PET/CT following admission, followed by CMR as described previously (Supplement text) (11).

#### Atherosclerotic disease by CTCA

Presence of coronary artery disease in each major coronary artery, and the main side branches were classified as potentially obstructive (>50% stenosis) or non-obstructive.

#### Myocardial disease by CMR

Ejection fraction (EF), regional wall motion abnormalities, myocardial fibrosis, oedema and presence of infarction in the left (LV) and right ventricles (RV) by late gadolinium enhancement (LGE) were determined as previously described (11). The anatomical 17-segment model was used to derive T1, T2 and extracellular volume (ECV) values for each segment excluding the apex (13). Acute myocardial inflammation was defined using the 2018 Lake Louise Criteria II that requires evidence of both myocardial oedema (high T2) and non-ischaemic injury (high T1, high ECV or non-ischemic LGE) (14). A sensitivity analysis was presented using the more sensitive criteria requiring the presence of abnormally high T1 *or* T2 values in conjunction with evidence of pericarditis or systolic dysfunction.

#### Myocardial, vascular and pulmonary pathology by 18F-FDG-PET/CT

As previously described (5) CT and 18F-FDG-PET scan images were co-registered and analysis performed using the 17-segment anatomical framework (13). Myocardial uptake was scored based on a visual scale. Patients with focal or diffuse uptake were identified as having acute myocardial inflammation (5).

Semi-quantative vascular inflammation on 18F-FDG-PET/CT for the aorta was assessed by the American Society of Nuclear Cardiologists visual grading criteria (15). Quantitative assessment was also undertaken on large vessel inflammation (6). In brief, a maximum arterial standardised uptake value (SUV) was derived in serial axial measurements across the ascending, arch and descending aorta. The target-to-background ratio (TBR) for each aortic region was calculated by averaging the ratio of maximum arterial SUV to mean venous SUV for each segment. Twenty-one age- and sex-matched historical controls who had previously undergone clinical 18F-FDG-PET/CT scans for other indications (eg. investigation of pulmonary nodules and reported as normal) and five healthy active controls were also scanned.

For pulmonary analysis, chest CT and 18F-FDG-PET/CT images were analysed separately for lung consolidation and inflammation respectively. Three-dimensional lung contours were generated and linked to the co-registered PET and CT images. Thresholds, for pathology, were determined at three pooled standard deviations above the population means. Control patients were used to define thresholds to delineate consolidation on CT (by lung density in Hounsfield units) and inflammation on 18F-FDG-PET (by SUV). The volumes of consolidated lung and inflamed lung were presented as percentage of total lung volume.

### Statistical analysis

Baseline clinical and imaging data were expressed as the median (interquatile range) for continuous data and categorical data as proportions. Clinical and imaging data were presented by tertile of cardiac troponin (a priori analysis), presence of myocarditis on CMR, myocardial cell infiltration on PET and degree of pulmonary inflammation/consolidation. A priori hypothesis testing was carried out across categorical and continuous covariates by tertile of cardiac troponin (11). Exploratory hypothesis testing was further conducted when comparing clinical and imaging parameters by myocarditis and myocardial cell infiltration status. A p-value of <0.05 was considered as statistically significant. No correction for multiple testing was done. Analysis was done in R (Version 4.0.3 http://www.R-project.org/).

## RESULTS

### Study Population

Of sixty-four consecutive patients with acute COVID-19, 33 were recruited (median age: 51 years [IQR 34, 55], 31 (94%) male and 31 (94%) of black ethnicity, Table 1). Thirteen patients declined to participate, and 18 had exclusion criteria. Twenty-four (73%) were hospitalised due to respiratory symptoms of cough with or without shortness of breath in the context of COVID-19. The remaining had non specific viral symptoms (fever, myalgia, arthralgia fatigue, diarrhoea, nausea or vomiting). No patients had been vaccinated. Twenty-nine patients underwent cardiac 18F-FDG-PET/CT, 26 CTCA and 26 CMR scanning (Supplemental Figure 1). CTCA and 18F-FDG-PET/CT scans were performed at a median time of four days after presentation (IQR 2-9 days). CMR scans were performed at a median time of 10 days (IQR 5-20 days). Six COVID-19 negative patients were recruited as controls.

**Table 1:**
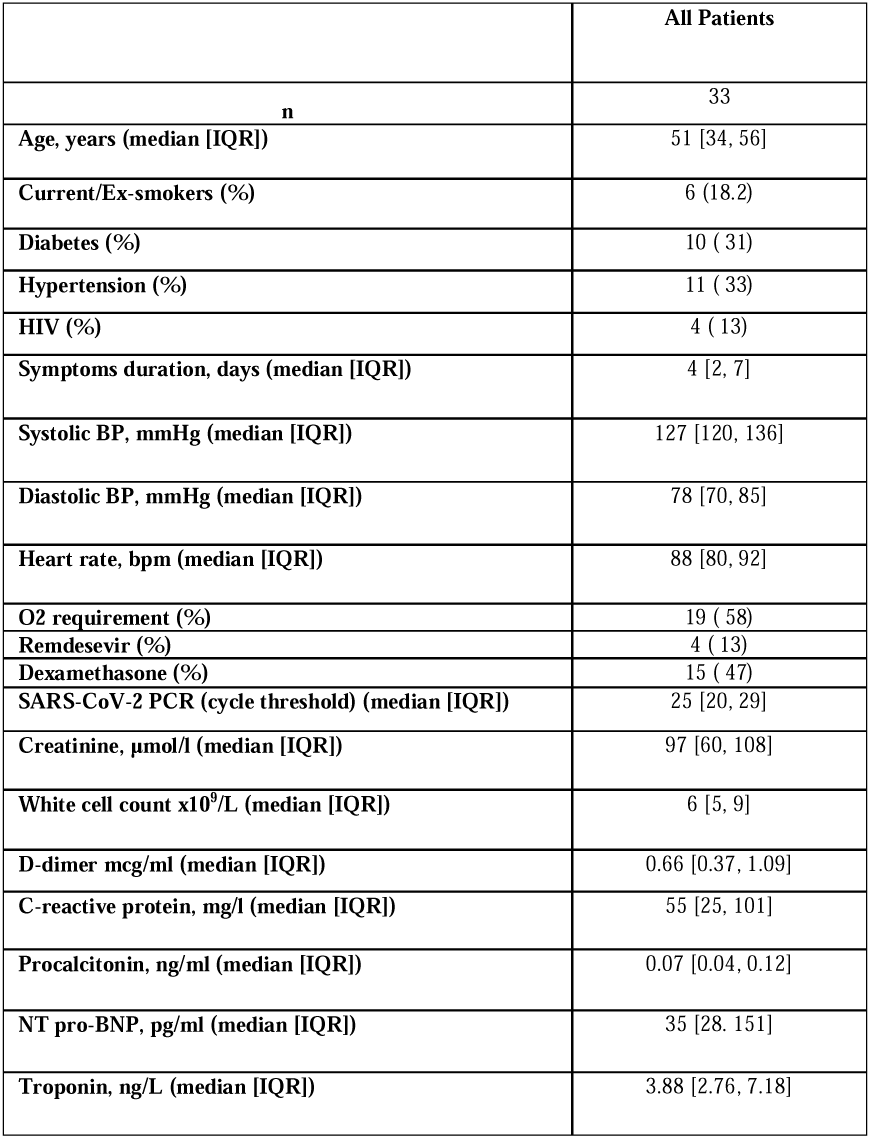
Baseline characteristics of patients with acute COVID-19.

|  | **All Patients** |
| --- | --- |
| **n** | 33 |
| **Age, years (median [IQR])** | 51 [34, 56] |
| **Current/Ex-smokers (%)** | 6 (18.2) |
| **Diabetes (%)** | 10 ( 31) |
| **Hypertension (%)** | 11 ( 33) |
| **HIV (%)** | 4 ( 13) |
| **Symptoms duration, days (median [IQR])** | 4 [2, 7] |
| **Systolic BP, mmHg (median [IQR])** | 127 [120, 136] |
| **Diastolic BP, mmHg (median [IQR])** | 78 [70, 85] |
| **Heart rate, bpm (median [IQR])** | 88 [80, 92] |
| **O2 requirement (%)** | 19 ( 58) |
| **Remdesevir (%)** | 4 ( 13) |
| **Dexamethasone (%)** | 15 ( 47) |
| **SARS-CoV-2 PCR (cycle threshold) (median [IQR])** | 25 [20, 29] |
| **Creatinine, $\mu\text{mol/l}$ (median [IQR])** | 97 [60, 108] |
| **White cell count $\times 10^9/\text{L}$ (median [IQR])** | 6 [5, 9] |
| **D-dimer mcg/ml (median [IQR])** | 0.66 [0.37, 1.09] |
| **C-reactive protein, mg/l (median [IQR])** | 55 [25, 101] |
| **Procalcitonin, ng/ml (median [IQR])** | 0.07 [0.04, 0.12] |
| **NT pro-BNP, pg/ml (median [IQR])** | 35 [28, 151] |
| **Troponin, ng/L (median [IQR])** | 3.88 [2.76, 7.18] |

The prevalence of biochemical evidence of myocardial injury (hsTrop >99^th^ centile upper reference limit) was 5% (n=2/31). Tertiles of hsTrop levels only correlated with CRP (22mg/L [IQR 12-32] vs 85 [50-100] vs 153[59-194], p=0.001) (Supplemental table 2). 25 patients underwent assessment of viral load by cycle threshold testing (7 high, 19 medium, 5 low) (Supplemental table 3). There was no association of viral load by cycle threshold (25 [IQR 25-28] vs 27 [IQR 22-29] vs p=0.57), CRP (34 mg/L [IQR 13-75] vs 45 mg/L [IQR 30-101], p=0.47), NT-proBNP (35 pg/ml [IQR 9-252] vs 35 pg/ml [IQR 28-58], p=0.89) or procalcitonin (0.04 ng/ml [IQR 0.02–0.08] vs 0.11 ng/ml [IQR 0.05-0.12], p=0.17) levels comparing patients with and without myocarditis.

### Cardiac magnetic resonance imaging

Twenty-six patients underwent CMR scanning. All scans were of adequate quality for volume and wall motion analysis. One scan was of insufficient quality for T1 mapping analysis, one insufficient quality for T2 analysis and one scan inadequate for LGE analysis. Myocarditis status was therefore available in 25 patients using the specific 2018 Lake Louise Criteria (14).

In the patient population the median LV EF was 51% [IQR 57-57] and RV EF 55% [48-50]. Median global native T1 was 1275ms [IQR 1250-1317], global ECV was 25% [24-28] and global T2 was 51ms [47-54]. Nine patients (35%, n=9/25) had LGE. Of these, two (22%) had subendocardial LGE, and eight (89%) mid-wall or epicardial LGE. Of these nine patients with LGE, seven (78%) also had evidence of active myocardial oedema by T2 value.

Nine (35%, n=9/25) patients had evidence of active myocarditis by the most specific 2018 Lake Louise criteria (Table 2, Figure 1). Six (57%, n=5/9) of these patients had evidence of LGE, with four in a myocarditis pattern (mid-wall), one subendocardial and one both. Thirteen patients (50%, n=13/25) had evidence of myocarditis by the sensitive criteria (Supplemental Table 4).

**Table 2:** Cardiac MRI results stratified by the specific 2018 Lake Louis II diagnosis of myocarditis, cardiac 18F-FDG-PET/CT evidence of myocardial inflammatory cell infiltration and presence of pulmonary inflammation or consolidation. *Denominators differ for each modality as not all scans are performed/diagnostic on every patient

|  | All Patients | Myocarditis (CMR) | No Myocarditis (CMR) | Inflammatory cell infiltration (Cardiac PET) | No Inflammatory cell infiltration (Cardiac PET) | <25 %pulmonary inflammation (PET) | ≥25 % pulmonary inflammation (PET) | <15% Pulmonary Consolidation (CT) | ≥15% Pulmonary Consolidation (CT) |
| --- | --- | --- | --- | --- | --- | --- | --- | --- | --- |
| **n** | 26* | 9 | 16 | 7 | 16 | 9 | 16 | 10 | 15 |
| **LV Ejection fraction, % (median (IQR))** | 61 [57, 67] | 59 [56, 62] | 64 [59, 68] | 62 [60, 63] | 60 [56, 69] | 57 [55, 62] | 64 [60, 68] | 58 [54, 64] | 64 [60, 68] |
| **LV EDVi, ml/m2 (median (IQR))** | 65 [62, 72] | 65 [60, 73] | 66 [62, 72] | 64 [56, 75] | 66 [62, 72] | 66 [60, 73] | 65 [64, 72] | 64 [60, 72] | 65 [64, 76] |
| **LV ESVi, ml/m2 (median (IQR))** | 25 [21, 29] | 25 [21, 32] | 22 [21, 27] | 22 [20, 32] | 26 [21, 30] | 26 [21, 32] | 22 [21, 27] | 26 [22, 30] | 22 [20, 28] |
| **LV SVi, ml/m2 (median (IQR))** | 42 [34, 48] | 38 [30, 44] | 44 [38, 49] | 38 [31, 42] | 44 [36, 50] | 41 [32, 46] | 44 [37, 49] | 38 [32, 48] | 44 [38, 48] |
| **RV Ejection fraction, % (median (IQR))** | 56 [48, 60] | 50 [47, 58] | 58 [54, 60] | 52 [48, 56] | 59 [52, 61] | 54 [45, 59] | 56 [52, 60] | 54 [49, 59] | 57 [51, 60] |
| **T1 - max, ms (median (IQR))** | 1356 [1304, 1412] | 1342 [1301, 1403] | 1372 [1322, 1414] | 1317 [1297, 1405] | 1380 [1335, 1424] | 1407 [1369, 1415] | 1321 [1298, 1400] | 1376 [1337, 1412] | 1334 [1299, 1424] |
| **T2 - max, ms (median (IQR))** | 62 [57, 68] | 68 [67, 70] | 58 [56, 62] | 66 [64, 69] | 60 [56, 63] | 62 [57, 67] | 62 [56, 67] | 60 [57, 67] | 62 [58, 67] |
| **ECV, max % (median (IQR))** | 31 [28, 34] | 34 [31, 36] | 29 [27, 32] | 33 [31, 35] | 30 [27, 34] | 32 [26, 34] | 30 [29, 34] | 30 [26, 34] | 31 [29, 35] |
| **LGE (%)** | 9 (35) | 6 ( 67) | 2 (12) | 6 ( 86) | 3 (19) | 3 (33) | 6 (38) | 4 (40) | 5 (33) |
| **Sub-endocardial LGE (%)** | 2 ( 8) | 2 ( 22) | 0 ( 0) | 2 ( 29) | 0 ( 0) | 0 ( 0) | 2 (12) | 0 ( 0) | 2 (13) |
| **Mid-wall LGE (%)** | 8 (31) | 5 ( 56) | 2 (12) | 5 ( 71) | 3 (19) | 3 (33) | 5 (31) | 4 (40) | 4 (27) |
| **Myocarditis (CMR) Specific Lake Louis criteria - n (%)** | 9 (36) |  |  | 5 ( 71) | 3 (20) | 3 (33) | 5 (33) | 3 (33) | 5 (33) |
| **Myocardial inflammatory cell infiltration (PET) - n (%)** | 7 (30) | 5 ( 63) | 2 (14) |  |  | 1 (11) | 6 (43) | 1 (11) | 6 (43) |
| **Pulmonary Inflammation - % of lung (median (IQR))** | 17 [3, 29] | 16 [2, 30] | 17 [10, 31] | 7 [2, 20] | 22 [13, 38] | 38 [29, 42] | 10 [1, 17] | 35 [29, 41] | 7 [1, 15] |
| **Pulmonary consolidation - % of lung (median (IQR))** | 12 [7, 18] | 10 [8, 16] | 13 [7, 18] | 9 [8, 12] | 16 [7, 19] | 18 [17, 26] | 8 [6, 13] | 18 [18, 25] | 8 [5, 11] |

**Figure 1:**
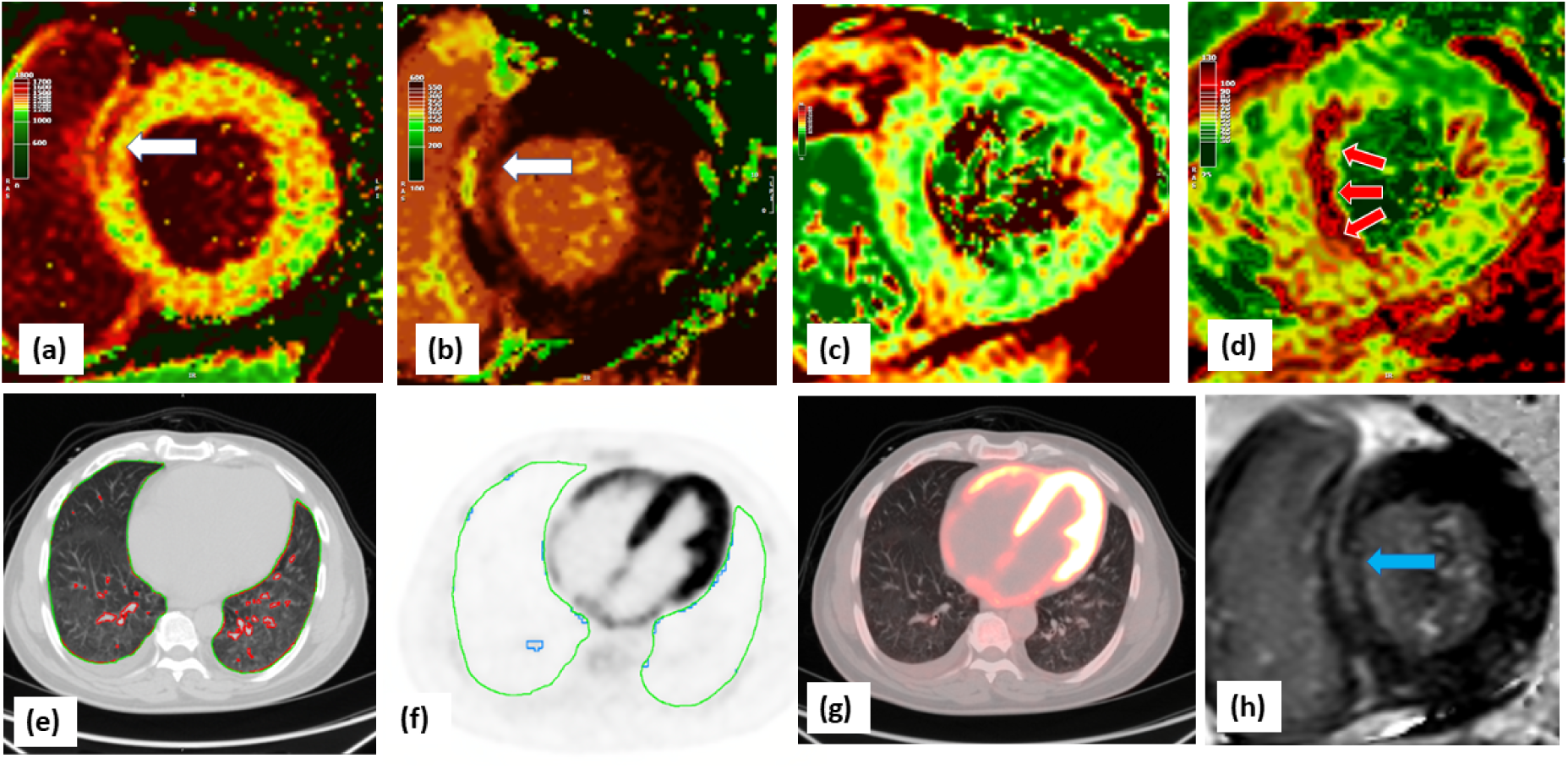
Severe myocarditis with minimal lung injury. There is mid-wall injury at the basal myocardium in the septum (white arrows) shown by CMR (a) native T1, (b) post contrast T1 and LGE (h, blue arrow). There was no increase in T2 values in this basal region (c), but there is gross increase in mid-ventricular septal T2 (d, red arrows) indicating oedema remote to prior myocardial fibrosis. There was minimal lung consolidation (e, red contours) or inflammation (f, blue contours). There is diffuse bi-ventricular 18F-FDG uptake (significantly higher than in the liver) (g). The patient had severe left and right ventricular impairment with elevated hsTrop (110 ng/L) and NT-proBNP (7140 pg/ml) but low CRP (10 mg/L).

Cardiac troponin concentrations were numerically higher in patients with myocarditis compared to those without (8.4 [IQR 4.0-55.3] vs 3.5 [2.5-5.5], p=0.07) (Supplemental Table 5). No differences in viral load (25 [IQR 20-28] vs 27[22-29], p=0.70), LV diastolic volume (55 ml/m2 [50-73] vs 55 [52-72], p=0.84) or LV ejection fraction (59% [55-52] vs 54 [59-58], p=0.23) were found in patients with and without myocarditis (Table 1, Supplemental Table 1 & Table 2).

### Computed-tomography coronary angiography

Twenty-five patients underwent CTCA and all had sufficient image quality. No patients had significant obstructive coronary artery disease (lumen stenosis>50%, Figure 2).

**Figure 2:**
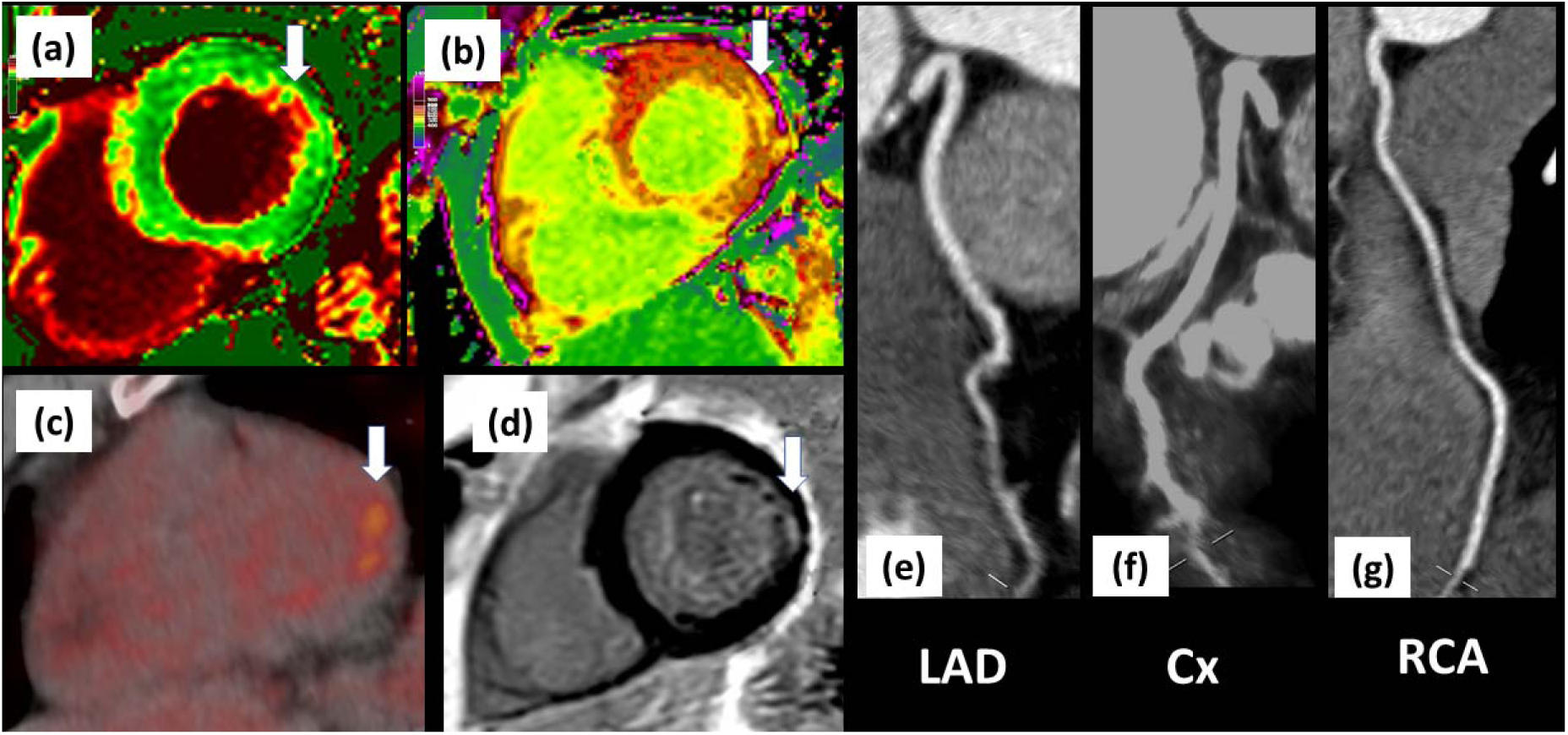
Focal infero-lateral myocarditis with no atherosclerotic disease. Changes (white arrows) in the native and post contrast CMR T1 values (a & b), 18F-FDG PET focal uptake (c) and sub-endocardial fibrosis on CMR late gadolinium enhancement (d). There was no significant coronary artery disease on CTCA (e, f & g). Biochemical cardiac and inflammatory markers were low (hsTrop 2.72 ng/L, NT-proBNP <35 pg/ml, CRP 4 mg/L).

### Positron emission tomography/computed-tomography

#### Vascular inflammation

Arterial inflammation in the ascending aorta by TBR was 1.97±0.35 (Supplemental Figure 2) and similar to historical or active controls (1.92 ± 0.32 and 2.03±0.05, p=0.74). There was no significant regional variation in TBR values in different aortic segments (Supplemental Table 7). No patients fulfilled the visual criteria for inflammation in the aorta or carotids (Supplemental Figure 3). There was no correlation with aortic FDG uptake (TBR) and CRP, hsTrop or viral load (Supplemental Table 8). Ascending aorta TBR was similar in patients with and without CMR myocarditis (1.93±0.18 vs 2.00±0.44, p=0.55) and with and without myocardial inflammatory cell infiltration on 18F-FDG-PET (1.97±0.17 vs 1.91±0.21, p=0.47).

#### Myocardial inflammatory cell infiltration

Two patients were not adequately fasted for cardiac analysis. Of the remaining, eight (30%, n=8/27) patients had evidence of myocardial inflammatory cell infiltration. Three patients had focal uptake (Figure 2), four focal on diffuse (Figure 3) and one diffuse (Figure 1).

**Figure 3:**
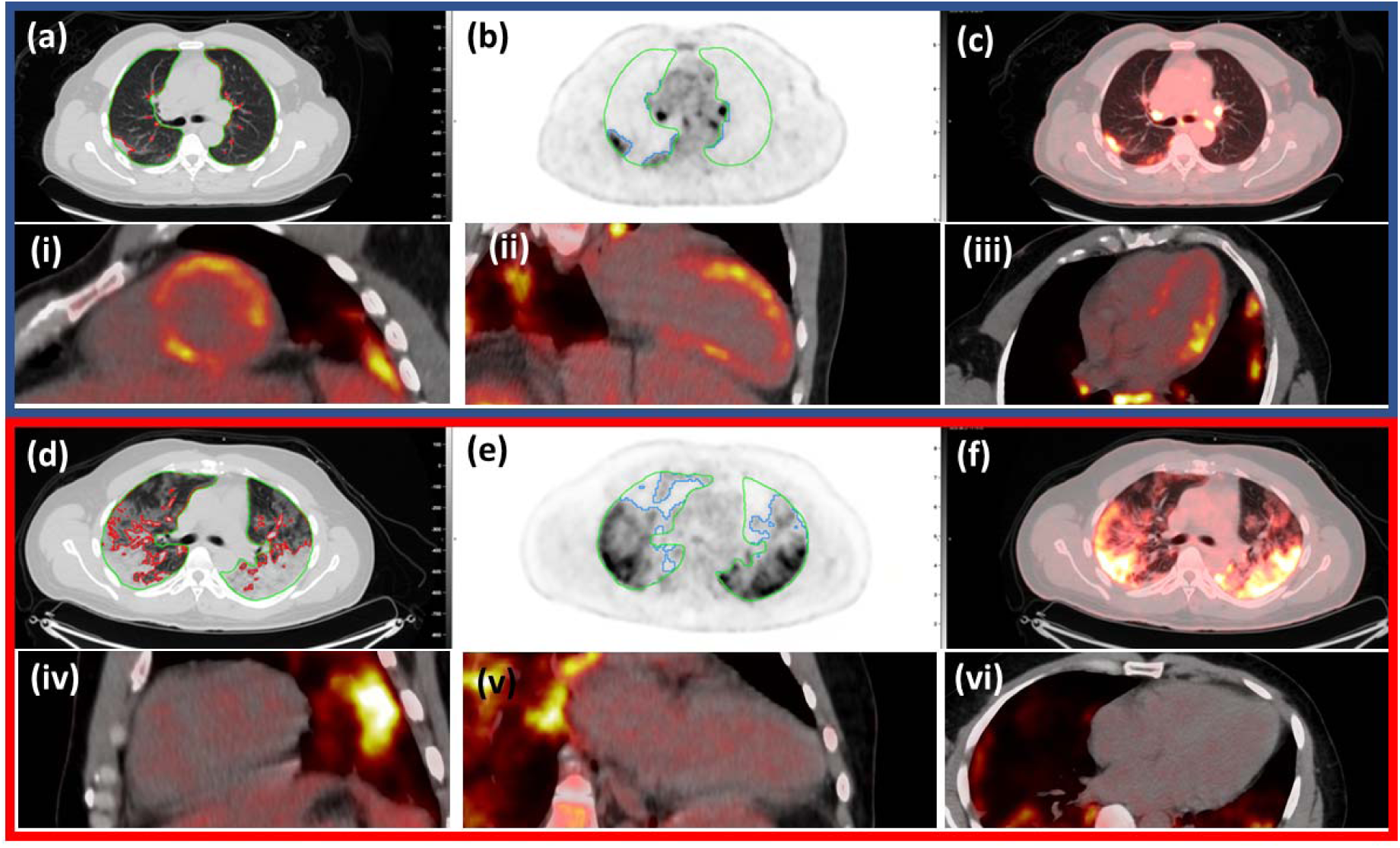
Cardiac and pulmonary 18F-FDG PET/CT imaging in 2 patients showing discordance between pulmonary and myocardial involvement. Top panel (blue outline) represents a patient with significant myocardial inflammatory cell infiltration with some pulmonary involvement - 17% lung consolidation and 29% inflammation. Cardiac inflammatory cell infiltration (focal on diffuse bright spots in lateral anterior and septal walls). Bottom panel (red outline) represents another patient with no myocardial involvement but with significant lung consolidation (35%) and inflammation (54%). Lung consolidation on CT (a & d, red contours), lung inflammation (b & e, blue contours), green contours lung parenchyma. Fused image (c & f showing lung inflammation with heat map on CT. Cardiac 18F-FDG PET shows inflammatory cell infiltration in the short axis (i & iv), vertical long-axis (ii & v) and horizontal long-axis views (iii & vi).

Twenty-two patients had both CMR and 18F-FDG-PET/CT. Of these eight patients had CMR-defined myocarditis by using the specific 2018 Lake Louise criteria, and 5/8 (53%) also had simultaneous evidence of inflammatory cell infiltration. Similarly, of the eight patients who had evidence inflammatory cell infiltration by 18F-FDG-PET/CT, 5/8 (53%) had myocardial oedema on CMR. 12 patients (55%, n=12/22) had no evidence of myocarditis or cellular uptake on CMR or 18F-FDG-PET/CT.

#### Pulmonary injury

Twenty-nine patients with acute COVID-19 infection, and five controls underwent pulmonary 18F-FDG-PET/CT. Overall, 25 patients had both CMR and pulmonary 18F-FDG-PET/CT performed.

In patients with acute COVID-19 the median amount of inflammation (based on 18F-FDG-PET) and consolidation (based on CT) as a percentage of total lung volume was 17% (IQR 5, 31) and 11% (IQR 7, 18) respectively. In controls, there was 0.18% (IQR 0.15, 0.57) inflammation and 3.0% (IQR 2.7, 3.1) consolidation.

When categorising patients who underwent both CMR and pulmonary 18F-FDG-PET/CT into tertiles, 7/25 patients had 0-5%, 9/25 had 5-25% and 9/25 had >25% inflammation of the total lung volume (Supplemental Table 7). Similarly, 5/25 patients had 0-7%, 10/25 had 7-15% and 10/25 had >15% consolidation of total lung volume. (Supplemental Table 9).

The degree of lung inflammation (15% [IQR 2-30%] vs 17% [10-31%], p=0.95) or consolidation (10% [8-15%] vs 13% [7-18%], p=0.85) was comparable in patients with and without myocarditis (Figure 3). Similarly, the degree of lung inflammation (7% [IQR 2-20%] vs 22% [13-38%], p=0.11) or consolidation (9% [8-12%] vs 15% [7-19%], p=0.23) was comparable in patients with and without myocardial inflammatory cell infiltration.

There was no association between the presence of myocarditis and the degree of lung injury. Of patient with CMR-based myocarditis 3/9 (37.5%) had severe pulmonary inflammation, compared to 5/17 (35.3%) without myocarditis (p=1.0). Patients with myocardial inflammatory cell infiltration, compared to those without, has numerically lower pulmonary inflammatory involvement (5% [2%-17%] versus 24%[13%-38%], p=0.05). There was no correlation between severity of lung inflammation (55% [IQR 52-50%] vs 54% [45-59], p=0.57) or consolidation (57% [51-50] vs 54% [49-59], p=0.52) with right ventricular (RV) ejection fraction. Similarly, no correlation was seen for severity of lung inflammation (81 ms/m2 [IQR 71-85%] vs 71 [55-79], p=0.35) or consolidation (79 ms/m2 [70-85] vs 74 [55-82], p=0.52) with indexed RV diastolic volumes (Table 2).

## DISCUSSION

To our knowledge this is the first study to systematically use molecular imaging, alongside anatomical and functional modalities, to explore cardiovascular and pulmonary pathobiology in acute COVID-19 infection. We make some important observations. First, rates of myocarditis (by CMR criteria) and myocardial inflammatory cell infiltration (by 18F-FDG-PET/CT imaging) were significant at 35% and 30% respectively. Second, the median burden of lung inflammation and consolidation was quantified at 17% and 11% of total lung volume respectively. Lung involvement, both inflammation and consolidation, did not correlate with presence of myocarditis or myocardial inflammatory cell infiltration. Third, vasculitis was not present in acute COVID-19. Finally, biochemical evidence of myocardial injury was not common with only two acute COVID-19 patients showing elevated troponin levels.

Our rates of myocarditis, despite recruiting patients with acute COVID-19, were lower than previously reported (1) but similar to other recent studies (2, 3). This in part reflects our choice of using the more specific 2018 Lake Louise criteria to define CMR-based myocarditis. Indeed, the prevalence of myocarditis rose from 1 in 3, to 1 in 2 when applying the most sensitive criteria as in previous studies (1). Using 18F-FDG-PET/CT imaging myocardial inflammatory cell infiltration was present in 1 in 3 cases. Surprisingly, neither the presence of myocarditis nor myocardial cell infiltration was associated with biochemical evidence of cardiac injury. Myocarditis may not always result in cell necrosis and troponin release (16, 17). Further, troponin release my be dynamic (18, 19) and may not be appreciated on single point blood sampling on hospital admission. Finally, studies on myocarditis have generally been restricted to patients with troponin elevations in whom significant coronary disease has been excluded (20). In contrast, our study involved cardiac imaging of an unselected population with an acute viral infection, regardless of troponin concentration.

Whilst CMR-based tissue characterisation can indicate myocardial oedema, molecular imaging with 18F-FDG-PET/CT reflects myocardial cellular infiltration - a better indicator of an acute inflammatory process (21, 22). Of those patients who had CMR defined myocarditis, only 53% had inflammatory cell presence. This suggests acute myocardial inflammation may have either occurred prior to presentation, or oedema is not always due to direct cellular infiltration. SARS-CoV-2 infection is present in the myocardium in the majority of individuals dying from COVID-19 (22). Furthermore, *in vitro* studies have shown SARS-CoV-2 cytopathic infection of cardiac myocytes with macrophage and lymphocytic infiltration (1, 22, 23). However, a cytokine storm has also been implicated in COVID-19 infection (24, 25). This process occurs sometime after viral inoculation, and may also result in cardiac pathology without presence of SARS-CoV-2 in the myocardium (26). In this case, systemic cytokines may also cause systemic capillary leak (with resultant oedema) without cellular infiltration of all tissues (27). Therefore cardiac injury may result from a dual injury process; initially from viral infection, followed by a subsequent cardiac insult from a systemic inflammatory response. In keeping with this, we demonstrated that some patients had evidence of prior myocardial fibrosis without associated oedema, but then also had active oedema without fibrosis in other regions (Figure 1).

Whilst the pathogenesis of hypercoagulability in COVID-19 remains unclear, vascular thrombosis has been described in hospitalised patients (28). Endothelial injury and vascular inflammation have been postulated to play a central role (9, 29, 30). In contrast, our study did not find any supporting evidence of arterial inflammation in acute COVID-19. We further found no evidence of coronary thrombosis to explain the myocardial pathology observed on imaging (Figure 2).

Previous studies had demonstrated coronary artery obstruction and ischaemic injury patterns on CMR, however the study population was restricted to those with troponin elevations (3). As such, we can conclude that the mechanism of cardiac pathology in acute COVID-19 infection is unlikely to have occurred secondary to coronary atherosclerosis, and the reported high prevalence of vascular thrombosis is not due to an arterial vasculitic process (28).

Macrophages and monocytes are known to be involved in the pathogenesis of acute respiratory distress syndrome, and there is growing evidence of their involvement in COVID-19 related pulmonary injury (31). We showed that the degree of pneumonitis, by 18F-FDG-PET/CT, was variable, correlated degree of lung consolidation but was not associated with presence of myocarditis. This suggests that myocarditis can occur in patients with minimal lung involvement.

Our study has some limitations. Firstly, although achieving comprehensive phenotyping this was an observational study in a small COVID-19 population. Almost half of the patients received either dexamethasone or remdesevir which may have supressed the inflammatory response and underestimated myocardial inflammation. Scanning, however, was performed early in the clinical course. Secondly, our assessment of vasculitis was based on 18F-FDG-PET/CT uptake in the large vessels. Vascular inflammation in the smaller vessels, due to limited spatial resolution, may be undetected. However, if vascular inflammation was secondary to a systemic cytokine storm or immune response, it would have been expected that this would have been reflected in the aorta and the medium sized carotids. Thirdly, we excluded patients with severe COVID-19 infection who were unable to tolerate imaging limiting the generalizability of our findings in this population. Finally, we did not perform cardiac biopsy. Although this is the gold standard for the diagnosis of myocarditis, we performed deep phenotyping using three different imaging modalities. The combination of myocardial inflammatory cell identifaction by 18F-FDG-PET, and myocarditis detection by CMR (using the strictest criteria to identify oedema), make our findings robust.

In conclusions, for the first time in acute COVID-19 infection and with the use of multi-modality imaging we make the following observations. Myocarditis was present in one in three patients and the majority of these patients had evidence of inflammatory cell infiltration by cardiac 18F-FDG-PET/CT. Pneumonitis was ubiquitous in acute COVID-19 but this inflammation was not associated with CMR myocarditis. The mechanism of cardiac pathology in acute COVID-19 is non-ischaemic, and vascular thrombosis in acute COVID-19 is not due to a vasculitic process that involves large or medium sized vessels.

## Data Availability

All data produced in the present study are available upon reasonable request to the authors

## ABBREVIATIONS

COVID-19: Coronavirus disease 2019
CTCA: Computed-tomography coronary angiography
18F-FDG-PET/CT: 2-deoxy-2-[fluorine-18]fluoro-D-glucose positron-emission tomography/computed-tomography
CMR: Cardiac magnetic-resonance
MISC-C: Multisystem inflammatory syndrome in children
hsTrop: High-sensitivity cardiac troponin I
NT-proBNP: N-Terminal pro B-type natriuretic peptide
CRP: C-Reactive protein
CTVL: Cycle threshold
EF: Ejection Fraction
LV: Left ventricle
RV: Right ventricle
LGE: Late gadolinium enhancement
ECV: Extracellular volume
SUV: Standardized uptake value
TBR: Target-to-background ratio
IQR: Interquartile range

## Acknowledgements

We would like to thank Rehema Sunday and Norah Matheka for coordinating the study.

## Trial registration

This study has been registered at the ISRCTN registry (ID ISRCTN12154994) on 14th August 2020. Accessible at www.isrctn.com/ISRCTN12154994

## Central Illustration

Patient with acute COVID-19 were scanned on hospital admission. CMR revealed myocarditis in 1 in 3 patients using the most stringest diagnostic criteria. A myocardial inflammatory cell infiltrate identified by 18F-FDG PET/CT was present in 30% of all patients, and in the majority of patients with CMR defined myocarditis. No patient had significant coronary artery disease on CTCA scanning. No patient had vasculitis. Although significant pulmonary inflammation and consolidation was common, it was not associated with the presence of myocarditis. Troponin testing did not identify patients with imaging evidence of myocardial oedema or inflammatory cell infiltration.

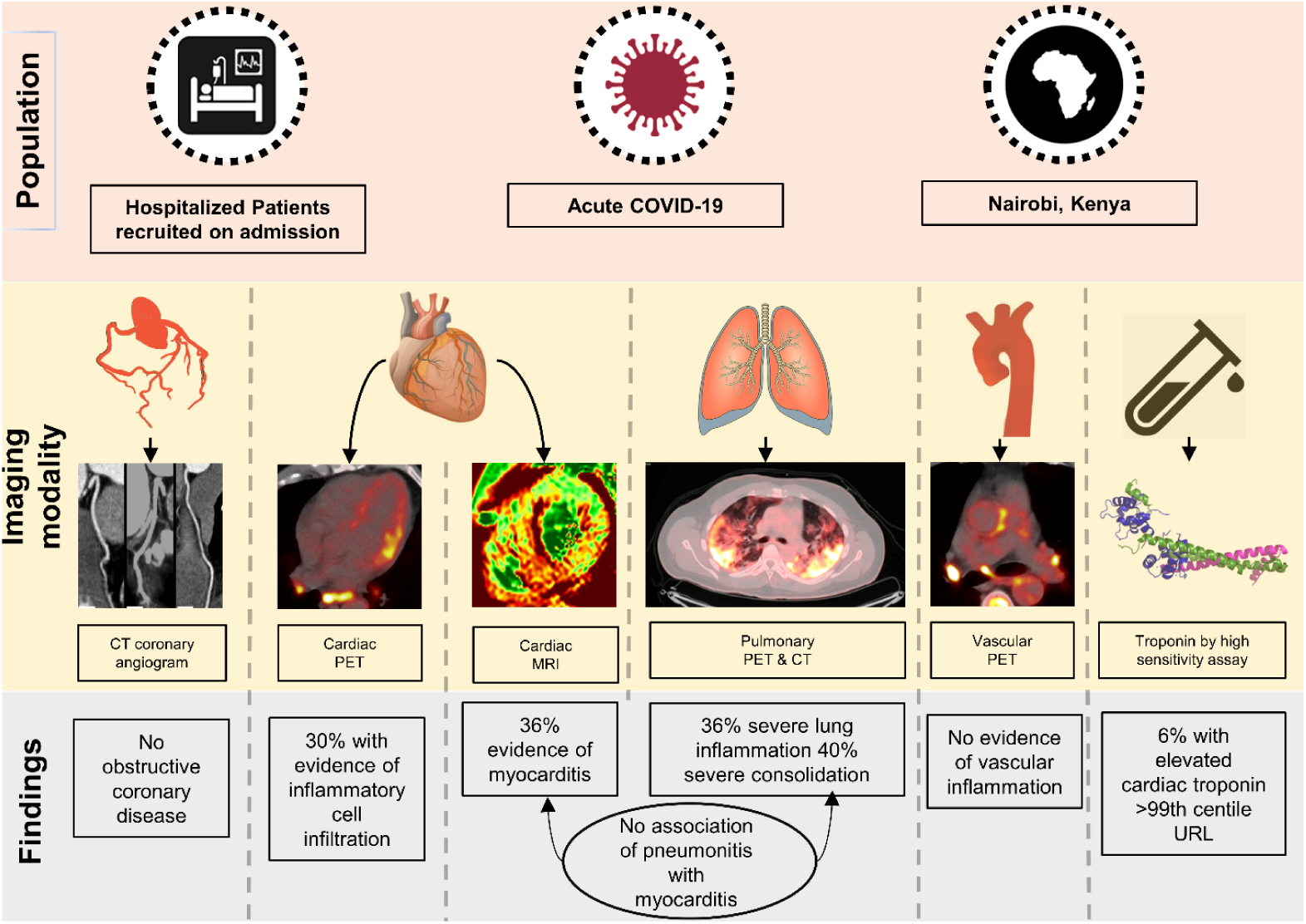

