## Supplement for "Assessment of cardiovascular & pulmonary pathobiology *in vivo* during acute COVID-19"

### **Assessment of cardiac, vascular & pulmonary pathobiology in vivo during acute COVID-19 – Supplementary Text**

**SCAN ANALYSIS**

**CMR**

CMR: CMR scans were analysed using dedicated software (Circle Cardiovascular Imaging Inc., Calgary, Canada). Control studies (5 participants; 80 segments) were used to determine T1, T2 and ECV cut-off values. In the controls, the mean LV EF was 62 ± 5 % and RV EF 61 ± 7 %. The median native T1, ECV and T2 across the segments was 1247ms (IQR 1225-1281), 27% (IQR 25-29) and 47ms (IQR 44-51) respectively. The 97.5 percentile used to identify abnormal segments on patient scans were 1384 ms for T1 and 64 ms for T2 relaxation times, and 31% for the ECV. No controls had subendocardial LGE and 1 had mid-wall LGE. The derived cut off values

For comparison, values over the 97.5 percentile of published normal values for T1 (1236 ms), T2 (64 ms) relaxation times and ECV (33%) for 3T CMR scanning were used.(1)

The myocardium was separated into 16 segments of the American Heart Association 17-segment model excluding the apex.(2) Manual endocardial and epicardial contours were drawn, and the segmentation was automated after identification of the superior RV insertion point. To ensure the blood pool or extra cardiac structures were excluded and only myocardium sampled, a 15% off-set was applied to both contours. T1 values ,T2 values, extra cellular volume and the presence of late gadolinium enhancement (LGE) were generated for each segment using dedicated software (Circle CVI).(3) T1 values indicated fibrosis or oedema, T2 values indicated oedema and gadolinium enhancement indicated the presence of infarction or fibrosis depending on distribution.(4-7)

Quantitative blinded analysis was performed by a trained consultant cardiologist with expertise in CMR (Manchester, UK).

**PET**

**Vascular**

18F-FDG-PET/CT scans were analysed using dedicated software (OsiriX 64-bit; OsiriX Imaging Software, Geneva, Switzerland). Semi qualitative vascular inflammation was assessed by the American Society of Nuclear Cardiologists visual grading criteria as follows: Grade 0 - No vascular uptake (≤ mediastinum), Grade 1: Vascular uptake < liver uptake, Grade 2: Vascular uptake = liver uptake, may be PET-positive, Grade 3: Vascular uptake > liver uptake, considered PET-positive.(8) Vascular inflammation was determined to be present in patients with Grade 2 or Grade 3 uptake.

Quantitative assessment was also undertaken large vessel inflammation.(9,10) In brief, regions of interest were drawn around the aorta in the axial position, repeated along the length of the aorta. A mean arterial SUV was derived from the average of the maximum SUV values in serial axial measurements across the whole aorta and in aortic segments (ascending, arch and the descending aorta). Similarly the average of mean SUV measurements from the venous pool derived the mean venous background SUV. The target-to-background (TBR) ratio was then calculated by dividing the maximum arterial SUV by the mean venous SUV. Twenty-one age and sex matched patients who had previously undergone clinical 18F-FDG-PET/CT scans and reported as normal (eg. investigation of pulmonary nodules) were used as historical controls. Five patients were also scanned as active controls.

Blinded analysis was performed by a trained consultant cardiologist with expertise in vascular PET scanning (Bristol, UK).

**Cardiac**

Standardised methodology for assessing myocardial inflammation PET/CT remains less well established. Myocardial uptake, on adequately fasted patients, was scored based on a visual scale and categorised as (i) none, (ii) focal uptake, (iii) focal on diffuse uptake, (iv) diffuse uptake (with uptake greater than the liver) or (v) non diagnostic (generalised uptake equal to or higherthan the liver). Liver SUV uptake was measured by drawing a hepatic region of interest. Patients with focal or diffuse uptake were identified as having acute myocardial inflammation. Visual uptake in the lateral myocardial wall was only identified as acute myocardial inflammation if uptake was >1.5 fold higher than in the septal or anterior walls.(11,12) Semi-qualitative blinded was performed by 2 consultant cardiologists and verified independently by a consultant cardio-thoracic radiologist specialised in nuclear radiology (Edinburgh & Manchester, UK and Nairobi, Kenya). Patients filled in a questionnaire before PET scanning, and were excluded from myocardial analysis if the fasting protocol was not adhered to.

**Pulmonary**

Chest CT and 18F-FDG-PET/CT images from hybrid scanner acquisitions were viewed and analyzed using MIM 7.1.2TM (MIM Software, Cleveland, OH). Three dimensional lung contours were first generated on CT using an automated density-based region-growing segmentation tool. Preliminary total lung contours were manually refined on each transaxial slice with a brush tool to include all well-aerated and consolidated lung tissue, while excluding proximal bronchovascular structures as well as mediastinal and hilar lymph nodes. Refined total lung contours were linked to the co-registered PET and CT images of all patients. In the control patient cohort, summary statistics of refined total lung contours (population mean, pooled standard deviation) were computed to define variation in CT-based normal lung density (in Hounsfield units [HU]) and PET-based physiologic background FDG uptake (in standardized uptake value [SUV]). Based on the control group summary statistics, thresholds to delineate consolidation on CT and inflammation on FDG-PET were set as 3 pooled standard deviations above the population mean HU and SUV, respectively. Within the refined total lung contours of the COVID-19 positive patient cohort, regions above the control group thresholds (-310 HU, 1.8 SUV) defined consolidated lung on CT and inflamed lung on FDG-PET. The volumes (absolute, relative fraction) of consolidated lung and inflamed lung were calculated. Examples of refined total lung, consolidated lung, and inflamed lung contours are shown in Figure 3.

Blinded analysis was performed by a trained nuclear radiologists with expertise in pulmonary PET scanning (Washington, USA).

**SUPPLEMENTARY TABLES**

|  | **CMR Myocarditis (specific 2018 Lake Louis criteria)** | **CMR No Myocarditis (specific 2018 Lake Louis criteria)** | **Inflammatory cell infiltration (Cardiac PET)** | **No Inflammatory cell infiltration (Cardiac PET)** | **Severe pulmonary inflammation (Lung PET)** | **Non-severe pulmonary inflammation (Lung PET)** | **Severe Pulmonary Consolidation** | **Non-Severe Pulmonary Consolidation** |
| --- | --- | --- | --- | --- | --- | --- | --- | --- |
| **n** | 9 | 17 | 8 | 19 | 11 | 18 | 11 | 18 |
| **Age, years (median [IQR])** | 51 [49, 59] | 48 [33, 52] | 50 [47, 54] | 51 [34, 57] | 52 [40, 59] | 50 [38, 54] | 52 [33, 59] | 51 [43, 56] |
| **Current or Ex-smokers (%)** | 2 ( 22) | 3 (18) | 1 ( 12) | 5 ( 26) | 3 (27) | 3 (17) | 2 (18) | 4 (22) |
| **Diabetes (%)** | 4 ( 50) | 3 ( 18) | 3 ( 43) | 5 ( 26) | 3 ( 27) | 5 ( 29) | 2 ( 18) | 6 ( 35) |
| **Hypertension (%)** | 4 ( 44) | 4 ( 24) | 2 ( 25) | 7 ( 37) | 3 ( 27.3) | 7 ( 38.9) | 3 ( 27) | 7 ( 39) |
| **HIV (%)** | 2 ( 29) | 1 ( 6) | 1 ( 17) | 0 ( 0) | 0 ( 0.0) | 2 ( 13) | 0 ( 0.0) | 2 ( 13) |
| **Symptoms duration, days (median [IQR])** | 5 [4, 7] | 4 [3, 7] | 5 [4, 6] | 3 [2, 7] | 4 [3, 8] | 3.50 [2, 7] | 7 [4, 8] | 3 [2, 5] |
| **Systolic BP, mmHg (median [IQR])** | 133 [125, 138] | 120 [120, 133] | 138 [124, 145] | 127 [115, 133] | 133 [119, 136] | 123 [120, 137] | 129 [124, 134] | 123 [120, 141] |
| **Diastolic BP, mmHg (median [IQR])** | 84 [81, 86] | 74 [70, 78] | 84 [80, 87] | 75 [70, 80] | 75 [72, 80] | 79 [70, 86] | 74 [70, 80] | 79 [71, 86] |
| **Heart rate, bpm (median [IQR])** | 86 [80, 90] | 90 [86, 100] | 88 [81, 93] | 88 [79, 90] | 89 [79, 93] | 87 [81, 92] | 90 [84, 103] | 86 [80, 90] |
| **O2 requirement (%)** | 6 ( 67) | 9 ( 52.9) | 4 ( 50) | 11 ( 58) | 10 ( 91) | 7 ( 39) | 9 ( 82) | 8 ( 44) |
| **Remdesevir (%)** | 1 ( 11) | 2 ( 13) | 0 ( 0) | 4 ( 22) | 4 ( 36) | 0 ( 0.0) | 4 ( 36) | 0 ( 0.0) |
| **Dexamethasone (%)** | 3 ( 33) | 9 ( 56) | 2 ( 25) | 10 ( 56) | 7 ( 64) | 7 ( 41) | 8 ( 73) | 6 ( 35) |
| **SARS-CoV-2 PCR (cycle threshold) (median [IQR])** | 26 [20, 28] | 27 [22, 29] | 26 [22, 27] | 25 [20, 29] | 23 [22, 29] | 27 [20, 29] | 26 [22, 29] | 26 [18, 29] |
| **Creatinine, μmol/l (median [IQR])** | 101 [82, 109] | 91 [79, 106] | 100 [83, 114] | 91 [78, 106] | 99 [78, 109] | 98 [82, 107] | 101 [78, 107] | 94 [80, 110] |
| **White cell count x10^9^/L (median [IQR])** | 6 [4, 7] | 6 [5, 9] | 6 [4, 9] | 6 [5, 8] | 7 [5, 9] | 6 [4, 8.] | 6 [4, 8] | 6 [5, 10] |
| **D-dimer mcg/ml (median [IQR])** | 0.21 [0.20, 0.52] | 0.88 [0.64, 1.32] | 0.58 [0.27, 2.42] | 0.84 [0.51, 1.29] | 0.88 [0.61, 1.39] | 0.70 [0.36, 0.94] | 0.66 [0.51, 0.88] | 0.83 [0.47, 1.31] |
| **C-reactive protein, mg/l (median [IQR])** | 34 [13, 75] | 68 [31, 101] | 16 [8, 43] | 96 [42, 148] | 124 [67, 153] | 42 [20, 82] | 101 [68, 151] | 36 [17, 82] |
| **Procalcitonin, ng/ml (median [IQR])** | 0.04 [0.02, 0.09] | 0.10 [0.05, 0.12] | 0.04 [0.03, 0.04] | 0.08 [0.05, 0.118] | 0.11 [0.05, 0.13] | 0.05 [0.04, 0.08] | 0.11 [0.08, 0.13] | 0.04 [0.04, 0.07] |
| **NT pro-BNP, pg/ml (median [IQR])** | 35 [9, 252] | 35 [28, 63] | 35 [28, 111] | 44 [32, 162] | 63 [25, 163] | 35 [35, 151] | 63 [31, 182] | 35 [35, 126] |
| **Troponin, ng/L (median [IQR])** | 8.41 [4.01, 55.35] | 3.51 [2.50, 5.58] | 4.44 [3.43, 8.34] | 3.62 [2.99, 7.19] | 5.58 [2.99, 8.41] | 3.62 [2.72, 6.89] | 4.60 [2.99, 6.88] | 4.14 [2.72, 7.20] |

**Supplemental Table 1:** Baseline characteristics of patients with acute COVID-19 stratified by CMR-defined myocarditis, cardiac 18F-FDG-PET/CT evidence of myocardial inflammatory cell infiltration and presence of pulmonary inflammation or consolidation.

|  | **Troponin by high sensitivity assay** | | |  |
| --- | --- | --- | --- | --- |
| **Tertile** | **1** | **2** | **3** |  |
| **Value (pg/L)** | **[2.50, 2.77)** | **[2.77, 6.59)** | **[6.59,2637.03]** | p |
| **n** | 8 | 13 | 11 |  |
| **Age, years (median [IQR])** | 46.50 [32.75, 51] | 49 [33, 52] | 56 [51, 59] | 0.05 |
| **Sex, male (%)** | 7 ( 87.5) | 13 (100.0) | 10 ( 90.9) | 0.45 |
| **Current / Exsmoker (%)** | 0 (0) | 2 (15.4) | 3 (27.3) | 0.27 |
| **Diabetes (%)** | 3 ( 42.9) | 3 ( 23.1) | 3 ( 27.3) | 0.64 |
| **Hypertension (%)** | 1 ( 12.5) | 3 ( 23.1) | 7 ( 63.6) | 0.04 |
| **Symptoms duration - days (median [IQR])** | 6.50 [5, 7] | 4 [3, 7] | 3 [2, 5.50] | 0.23 |
| **Systolic BP, mmHg (median [IQR])** | 120 [117.50, 123.75] | 129 [120, 135] | 133 [115, 138] | 0.36 |
| **Diastolic BP, mmHg (median [IQR])** | 76.50 [70, 83.25] | 79 [75, 88] | 70 [70, 84.50] | 0.59 |
| **Heart rate, bpm (median [IQR])** | 89 [87, 94] | 89 [81, 95] | 81 [79, 88] | 0.47 |
| **Oxygen requirement (%)** | 3 ( 37.5) | 6 ( 46.2) | 9 ( 81.8) | 0.1 |
| **Remdesevir (%)** | 0 ( 0.0) | 3 ( 25.0) | 1 ( 9.1) | 0.24 |
| **Dexamethasone (%)** | 2 ( 25.0) | 6 ( 50.0) | 7 ( 63.6) | 0.25 |
| **SARS-CoV-2 PCR (cycle threshold) (median [IQR])** | 14.96 [13.58, 26.12] | 27.94 [23.20, 29.23] | 24.40 [20.90, 30.38] | 0.09 |
| **Creatinine, μmol/l (median [IQR])** | 84 [74.50, 107.50] | 96 [79, 106.25] | 101 [92, 115] | 0.38 |
| **White cell count x10^9 (median [IQR])** | 5.48 [3.90, 7.43] | 5.77 [5.17, 8.60] | 6.79 [5.27, 9.25] | 0.52 |
| **Lymphocyte count x10^9 (median [IQR])** | 1.51 [1.22, 1.72] | 1.45 [1.23, 1.85] | 1.28 [1.00, 1.71] | 0.79 |
| **D-dimer mcg/ml (median [IQR])** | 0.93 [0.43, 1.56] | 0.65 [0.46, 0.80] | 0.74 [0.22, 1.04] | 0.87 |
| **C-reactive protein, mg/L (median [IQR])** | 22 [12, 32] | 86 [50, 100] | 153 [59, 194] | 0.001 |
| **Procalcitonin, ng/ml (median [IQR])** | 0.03 [0.01, 0.06] | 0.08 [0.04, 0.12] | 0.08 [0.05, 0.17] | 0.124 |
| **NT pro-BNP, pg/ml (median [IQR])** | 35 [20.49, 39.25] | 35 [28.12, 56.52] | 151 [58, 388.30] | 0.05 |
| **Inflammation, % of lungs (median [IQR])** | 15 [9, 22] | 22 [15, 38] | 21 [2, 32] | 0.55 |
| **Consolidation, % of lungs(median [IQR])** | 13 [9, 15] | 14 [7, 19] | 11 [8, 18] | 0.97 |
| **Myocarditis (MRI)** | 2 ( 28.6) | 1 ( 11.1) | 5 ( 55.6) | 0.13 |
| **Myocardial inflammatory cell infiltration (PET)** | 2 ( 33.3) | 3 ( 25.0) | 2 ( 20.0) | 0.84 |

**Supplemental Table 2:** Baseline characteristics stratified by troponin concentration.

|  | **Viral Load by Cycle Threshold** | | |
| --- | --- | --- | --- |
| **Viral load** | **High** | **Medium** | **Low** |
| **n** | 7 | 19 | 5 |
| **Age, years (median [IQR])** | 51 [46.50, 53.50] | 51 [37.50, 55.50] | 54 [33, 56] |
| **Sex, male (%)** | 6 ( 85.7) | 18 ( 94.7) | 5 (100.0) |
| **Current / Exsmoker (%)** | 1 (14.6) | 4 (11.1) | 1 (20) |
| **Diabetes (%)** | 5 ( 83.3) | 4 ( 21.1) | 1 ( 20.0) |
| **Hypertension (%)** | 3 ( 42.9) | 5 ( 26.3) | 3 ( 60.0) |
| **Symptoms duration - days (median [IQR])** | 5 [3, 7] | 5 [3, 7.50] | 2 [1, 5] |
| **Systolic BP, mmHg (median [IQR])** | 121 [120, 137] | 127 [120, 137] | 129 [110, 133] |
| **Diastolic BP, mmHg (median [IQR])** | 81 [78, 89] | 79 [70, 85] | 65 [64, 70] |
| **Heart rate, bpm (median [IQR])** | 88 [81, 89] | 90 [83, 99] | 80 [78, 89] |
| **Oxygen requirement (%)** | 1 ( 14.3) | 15 ( 78.9) | 3 ( 60.0) |
| **Remdesevir (%)** | 0 ( 0.0) | 3 ( 15.8) | 1 ( 20.0) |
| **Dexamethasone (%)** | 1 ( 14.3) | 11 ( 57.9) | 3 ( 60.0) |
| **SARS-CoV-2 PCR (cycle threshold) (median [IQR])** | 14.35 [13.55, 15.87] | 26.02 [22.46, 28.20] | 32.66 [32.28, 32.89] |
| **Creatinine, μmol/l (median [IQR])** | 88 [71, 114.50] | 96 [82.50, 107] | 106 [80, 106] |
| **White cell count x10^9 (median [IQR])** | 5.77 [4.73, 7.61] | 5.83 [4.53, 8.55] | 7.10 [5.19, 8.06] |
| **Lymphocyte count x10^9 (median [IQR])** | 1.65 [1.38, 1.78] | 1.25 [1.08, 1.46] | 1.31 [1.28, 1.75] |
| **D-dimer mcg/ml (median [IQR])** | 0.67 [0.24, 1.17] | 0.65 [0.39, 0.84] | 0.94 [0.64, 1.40] |
| **C-reactive protein, mg/l (median [IQR])** | 25 [9, 55.50] | 55 [38, 101] | 125 [74.50, 221.75] |
| **Procalcitonin, ng/ml (median [IQR])** | 0.06 [0.02, 0.07] | 0.08 [0.04, 0.12] | 0.16 [0.06, 0.37] |
| **NT pro-BNP, pg/ml (median [IQR])** | 35 [22.20, 43.50] | 35 [27.32, 84.50] | 189 [184, 252] |
| **Troponin, ng/L (median [IQR])** | 2.50 [2.50, 4.07] | 4.52 [3.15, 7.19] | 6.89 [3.62, 9.66] |
| **Inflammation, % of lungs (median [IQR])** | 0.15 [0.07, 0.17] | 0.26 [0.13, 0.39] | 0.18 [0.13, 0.26] |
| **Consolidation, % of lungs(median [IQR])** | 0.09 [0.07, 0.10] | 0.17 [0.08, 0.18] | 0.12 [0.11, 0.16] |
| **Myocarditis (MRI)** | 2 ( 50.0) | 6 ( 37.5) | 1 ( 20.0) |
| **Myocardial inflammatory cell infiltration (PET)** | 1 ( 20.0) | 6 ( 35.3) | 0 ( 0.0) |

**Supplemental Table 3:** Baseline characteristics stratified by viral load by cycle threshold.

* Viral load determined by cycle threshold. A lower cycle threshold indicates a higher viral load.

|  | **CMR Myocarditis (specific 2018 Lake Louis criteria)** | **CMR No Myocarditis (specific 2018 Lake Louis criteria)** | **CMR Myocarditis (sensitive 2018 Lake Louis criteria)** | **CMR No Myocarditis (sensitive 2018 Lake Louis criteria)** |
| --- | --- | --- | --- | --- |
| **n** | 9 | 16 | 13 | 13 |
| **LV Ejection fraction, % (median (IQR))** | 59 [56, 62] | 64 [59, 68] | 59 [56, 62] | 65 [59, 69] |
| **LV EDVi, ml/m2 (median (IQR))** | 65 [60, 73] | 66 [62, 72] | 62 [60, 71] | 66 [65, 76] |
| **LV ESVi, ml/m2 (median (IQR))** | 25 [21, 32] | 22 [21, 27] | 25 [21, 29] | 22 [21, 28] |
| **LV SVi, ml/m2 (median (IQR))** | 38 [30, 44] | 44 [38, 49] | 36 [32, 41] | 45 [43, 49] |
| **LV mass, g/m2 (median (IQR))** | 62 [57, 67] | 55 [52, 58] | 61 [54, 67] | 56 [52, 59] |
| **RV Ejection fraction, % (median (IQR))** | 50 [47, 58] | 58 [54, 60] | 52 [47, 58] | 60 [53, 61] |
| **RV EDVi, ml/m2 (median (IQR))** | 79 [64, 83] | 72 [70, 86] | 71 [64, 83] | 76 [71, 88] |
| **RV ESVi, ml/m2 (median (IQR))** | 38 [33, 40] | 32 [28, 40] | 34 [32, 40] | 34 [28, 44] |
| **RV SVi, ml/m2 (median (IQR))** | 38 [29, 43] | 43 [38, 49] | 38 [30, 43] | 44 [41, 49] |
| **Global T1 - mean, ms (median (IQR))** | 1270 [1241, 1302] | 1279 [1264, 1321] | 1269 [1241, 1302] | 1276 [1259, 1322] |
| **T1 - max, ms (median (IQR))** | 1342 [1301, 1403] | 1372 [1322, 1414] | 1342 [1299, 1407] | 1369 [1325, 1415] |
| **Global T2 - mean, ms (median (IQR))** | 54 [53, 56] | 49 [47, 51] | 54 [52, 56] | 49 [47, 51] |
| **T2 - max, ms (median (IQR))** | 68 [67, 70] | 58 [56, 62] | 68 [64, 69] | 58 [55, 62] |
| **Global ECV, % (median (IQR))** | 26 [25, 28] | 25 [24, 27] | 26 [24, 28] | 25 [24, 26] |
| **ECV - max % (median (IQR))** | 34 [31, 36] | 29 [27, 32] | 33 [30, 34] | 29 [27, 32] |
| **LGE present – n (%)** | 6 ( 67) | 2 (12) | 9 (69) | 0 (0) |
| **Sub-endocardial LGE present- n (%)** | 2 ( 22) | 0 ( 0) | 2 (15) | 0 ( 0.0) |
| **Mid-wall LGE present- n (%)** | 5 ( 56) | 2 (12) | 8 (62) | 0 ( 0.0) |
| **Myocardial inflammatory cell infiltration on PET - n (%)** | 5 ( 62) | 2 (14) | 7 (58) | 0 ( 0.0) |
| **Pulmonary Inflammation - % of lung (median (IQR))** | 16 [2, 30] | 17 [10, 31] | 22 (6, 30) | 17 (3, 26) |
| **Pulmonary consolidation - % of lung (median (IQR))** | 10 [8, 16] | 13 [7, 18] | 15 (9, 19) | 11 (7, 18) |

**Supplemental Table 4:** Cardiac and pulmonary imaging parameters stratified by specific and sensitive 2018 Lake Louis criteria for myocarditis.

|  | **Troponin** | | |  |
| --- | --- | --- | --- | --- |
|  | **2.50 - 2.77** | **2.77 - 6.59** | **>6.59** | **p** |
| **Tertile** | **1** | **2** | **3** |  |
| n | 7 | 9 | 9 |  |
| LV Ejection fraction, % (median (IQR)) | 60 [59, 64] | 59 [57, 66] | 65 [56, 69] | 0.696 |
| LV EDVi, ml/m2 (median (IQR)) | 64 [61, 70] | 65 [62, 71] | 66 [65, 71] | 0.906 |
| LV ESVi, ml/m2 (median (IQR)) | 25 [22, 26] | 27 [22, 29] | 21 [20, 25] | 0.314 |
| LV SVi, ml/m2 (median (IQR)) | 39 [36, 46] | 40 [34, 49] | 43 [30, 45] | 0.981 |
| LV mass, g/m2 (median (IQR)) | 54 [54, 60] | 57 [52, 69] | 57 [52, 61] | 0.911 |
| RV Ejection fraction, % (median (IQR)) | 53 [48, 58] | 54 [52, 59] | 58 [47, 61] | 0.797 |
| RV EDVi, ml/m2 (median (IQR)) | 71 [68, 81] | 83 [68, 88] | 71 [65, 79] | 0.475 |
| RV ESVi, ml/m2 (median (IQR)) | 34 [30, 40] | 36 [32, 47] | 33 [28, 38] | 0.575 |
| RV SVi, ml/m2 (median (IQR)) | 39 [36, 41] | 43 [37, 49] | 43 [29, 46] | 0.498 |
| Global T1, median (median (IQR)) | 1277 [1260, 1302] | 1281 [1248, 1306] | 1273 [1266, 1321] | 0.929 |
| T1, max (median (IQR)) | 1317 [1306, 1387] | 1403 [1335, 1414] | 1342 [1325, 1415] | 0.649 |
| Global T2, median (median (IQR)) | 53 [48, 54] | 48 [46, 49] | 53 [51, 60] | 0.013 |
| T2, max (median (IQR)) | 62 [60, 66] | 57 [56, 60] | 67 [62, 71] | 0.057 |
| Global ECV, median (median (IQR)) | 27 [24, 29] | 24 [23, 25] | 26 [25, 28] | 0.111 |
| ECV, max (median (IQR)) | 32 [31, 35] | 29 [27, 30] | 34 [28, 34] | 0.153 |
| Late gadolinium enhancement (%) | 1 ( 14) | 4 ( 44) | 3 ( 33) | 0.437 |
| Sub-endocardial LGE (%) | 1 ( 14) | 1 ( 11) | 0 ( 0) | 0.528 |
| Mid-wall LGE (%) | 0 ( 0) | 4 ( 44) | 3 ( 33) | 0.132 |
| Myocarditis (MRI) (%) | 2 ( 29) | 1 ( 12) | 5 ( 56) | 0.163 |
| Myocarditis (PET) (%) | 2 ( 40) | 2 ( 22) | 2 ( 25) | 0.762 |
| Pulmonary inflammation, % of lung (median(IQR)) | 15 [9, 22] | 26 [17, 38] | 13 [1, 32] | 0.513 |
| Pulmonary consolidated, % of lung (median(IQR)) | 13 [9, 15] | 17 [9, 18] | 12 [7, 18] | 0.882 |

**Supplemental Table 5:** Cardiac and pulmonary imaging parameters stratified by troponin results according to tertile.

|  | **Vascular 18F-FDG-PET/CT** | | | |
| --- | --- | --- | --- | --- |
|  | **Acute COVID-19** | **Active Controls** | **Historical Controls** | **p** |
| **n** | 29 | 5 | 21 |  |
| **Ascending aorta TBR, max SUV (mean (SD))** | 1.97 (0.35) | 2.03 (0.06) | 1.92 (0.32) | 0.744 |
| **Aortic arch TBR, max SUV (mean (SD))** | 2.00 (0.32) | 1.99 (0.15) | 1.92 (0.27) | 0.613 |
| **Descending aorta TBR, max SUV (mean (SD))** | 2.01 (0.44) | 1.85 (0.07) | 1.90 (0.59) | 0.644 |
| **Whole aorta TBR, max SUV (mean (SD))** | 2.01 (0.35) | 1.92 (0.08) | 1.91 (0.46) | 0.661 |

**Supplemental Table 6:** Vascular 18F-FDG-PET TBR by aortic region comparing acute COVID-19 cases to active and historical controls.

|  | **CRP (mg/L)** | | | **hsTroponin (ng/L)** | | | **Viral Load *** | | |
| --- | --- | --- | --- | --- | --- | --- | --- | --- | --- |
| **Tertile** | **1** | **2** | **3** | **1** | **2** | **3** | **Cut-off** | **Cut-off** | **Cut-off** |
| **Value** | **[4, 38]** | **[38,**  **100]** | **[100,**  **416]** | **[2.50, 2.77]** | **[2.77, 6.59]** | **[6.59,**  **2637]** | **Low VL**  **[CT >30]** | **Med VL**  **[CT 20-30]** | **High VL**  **[CT 20]** |
| **n** | 8 | 7 | 9 | 6 | 11 | 10 | 5 | 17 | 4 |
| **Ascending aorta TBR, max SUV (mean (SD))**** | 1.81 (0.12) | 1.93 (0.18) | 2.13 (0.54) | 1.88 (0.18) | 1.90 (0.17) | 2.09 (0.54) | 2.06 (0.81) | 1.99 (0.17) | 1.82 (0.12) |
| **Ascending aorta TBR, mean SUV (mean (SD))** | 1.32 (0.08) | 1.36 (0.12) | 1.44 (0.21) | 1.35 (0.14) | 1.39 (0.13) | 1.38 (0.20) | 1.39 (0.30) | 1.40 (0.11) | 1.27 (0.03) |
| **Aortic arch TBR, max SUV (mean (SD))** | 1.95 (0.30) | 2.08 (0.27) | 2.07 (0.42) | 1.90 (0.29) | 2.13 (0.26) | 1.91 (0.39) | 2.01 (0.54) | 2.06 (0.27) | 1.70 (0.14) |
| **Aortic arch TBR, mean suv (mean (SD))** | 1.31 (0.10) | 1.34 (0.13) | 1.39 (0.22) | 1.32 (0.16) | 1.37 (0.11) | 1.32 (0.21) | 1.35 (0.30) | 1.35 (0.12) | 1.23 (0.07) |
| **Descending aorta TBR, max SUV (mean (SD))** | 1.85 (0.27) | 2.13 (0.38) | 2.14 (0.64) | 1.93 (0.38) | 2.21 (0.55) | 1.85 (0.30) | 2.00 (0.55) | 2.08 (0.45) | 1.78 (0.37) |
| **Descending aorta TBR, mean SUV (mean (SD))** | 1.31 (0.12) | 1.43 (0.16) | 1.46 (0.24) | 1.39 (0.22) | 1.46 (0.18) | 1.32 (0.18) | 1.38 (0.20) | 1.41 (0.19) | 1.25 (0.16) |
| **Whole aorta TBR, max SUV (mean (SD))** | 1.88 (0.22) | 2.09 (0.27) | 2.12 (0.50) | 1.92 (0.28) | 2.15 (0.38) | 1.92 (0.35) | 2.02 (0.52) | 2.07 (0.33) | 1.76 (0.21) |
| **Whole aorta TBR, mean SUV (mean (SD))** | 1.31 (0.09) | 1.39 (0.13) | 1.43 (0.21) | 1.36 (0.17) | 1.42 (0.14) | 1.33 (0.18) | 1.37 (0.24) | 1.40 (0.14) | 1.25 (0.09) |

**Supplemental Table 7:** Vascular 18F-FDG-PET results stratified by CRP, hsTroponin and viral load. VL – Viral load. CT – Cycle Threshold

* Viral load determined by cycle threshold. A lower cycle threshold indicates a higher viral load.

** p-value for comparing TBR by tertile of CRP, hsTrop and Viral load were 0.20, 0.44 and 0.81 respectively

|  | **Lung inflammation (%)** | | |
| --- | --- | --- | --- |
|  | **0% - 4%** | **5% - 25%** | **>25%** |
| **n** | 7 | 9 | 9 |
| **LV Ejection fraction, % (median (IQR))** | 60 [60, 64] | 66 [64, 69] | 57 [55, 62] |
| **LV EDVi, ml/m2 (median (IQR))** | 66 [60, 76] | 65 [64, 71] | 66 [60, 73] |
| **LV ESVi, ml/m2 (median (IQR))** | 22 [20, 33] | 22 [21, 27] | 26 [21, 32] |
| **LV SVi, ml/m2 (median (IQR))** | 40 [32, 48] | 45 [39, 49] | 41 [32, 46] |
| **LV mass, g/m2 (median (IQR))** | 60 [58, 64] | 57 [53, 62] | 56 [48, 57] |
| **RV Ejection fraction, % (median (IQR))** | 58 [52, 61] | 55 [52, 60] | 54 [45, 59] |
| **RV EDVi, ml/m2 (median (IQR))** | 70 [64, 91] | 83 [74, 85] | 71 [65, 79] |
| **RV ESVi, ml/m2 (median (IQR))** | 33 [24, 42] | 36 [32, 40] | 33 [28, 47] |
| **RV SVi, ml/m2 (median (IQR))** | 41 [30, 52] | 44 [43, 49] | 38 [35, 43] |
| **Global T1, ms (median (IQR))** | 1244 [1242, 1293] | 1273 [1248, 1281] | 1306 [1291, 1323] |
| **T1 max, ms (median (IQR))** | 1334 [1299, 1422] | 1317 [1295, 1375] | 1407 [1369, 1415] |
| **Global T2, ms (median (IQR))** | 53 [51, 57] | 49 [47, 52] | 49 [48, 53] |
| **T2 max, ms (median (IQR))** | 66 [58, 70] | 60 [56, 63] | 62 [57, 67] |
| **Global ECV, % (median (IQR))** | 26 [25, 29] | 25 [23, 27] | 25 [24, 27] |
| **ECV max, % (median (IQR))** | 30 [29, 36] | 30 [27, 34] | 32 [26, 34] |
| **Late gadolinium enhancement, n (%)** | 3 (43) | 3 (33) | 3 (33) |
| **Sub-endocardial LGE, n (%)** | 0 (0) | 2 (22) | 0 (0) |
| **Midwall LGE, n (%)** | 3 (43) | 2 (22) | 3 (33) |
| **RV insertion point LGE, n (%)** | 2 (29) | 1 (11) | 1 (11) |
| **Oxygen requirement, n (%)** | 4 (57) | 3 (33) | 8 (89) |
| **Myocarditis (MRI) (%)** | 3 (43) | 2 (25) | 3 (33) |
| **Myocarditis (PET) (%)** | 3 (50) | 3 (38) | 1 (11) |
| **Pulmonary consolidated, % of lung (median(IQR))** | 7 [4, 8] | 10 [9, 16] | 18 [17, 26] |

**Supplemental Table 8:** Imaging parameters stratified by pulmonary inflammation

|  | **Lung Consolidation (%)** | | |
| --- | --- | --- | --- |
|  | **0% - 7%** | **7% - 15%** | **>15%** |
| **n** | 5 | 10 | 10 |
| **LV Ejection fraction, % (median (IQR))** | 60 [60, 69] | 64 [60, 68] | 58 [54, 64] |
| **LV EDVi, ml/m2 (median (IQR))** | 66 [65, 68] | 65 [64, 78] | 64 [60, 72] |
| **LV ESVi, ml/m2 (median (IQR))** | 22 [20, 28] | 22 [21, 27] | 26 [22, 30] |
| **LV SVi, ml/m2 (median (IQR))** | 45 [40, 52] | 44 [38, 46] | 38 [32, 48] |
| **LV mass, g/m2 (median (IQR))** | 59 [56, 60] | 59 [54, 62] | 56 [49, 62] |
| **RV Ejection fraction, % (median (IQR))** | 60 [60, 62] | 54 [48, 57] | 54 [49, 59] |
| **RV EDVi, ml/m2 (median (IQR))** | 70 [67, 74] | 84 [73, 87] | 74 [66, 82] |
| **RV ESVi, ml/m2 (median (IQR))** | 26 [21, 30] | 38 [33, 43] | 36 [29, 45] |
| **RV SVi, ml/m2 (median (IQR))** | 44 [41, 49] | 41 [39, 48] | 40 [36, 46] |
| **Global T1, ms (median (IQR))** | 1266 [1244, 1273] | 1273 [1244, 1311] | 1297 [1271, 1317] |
| **T1 max, ms (median (IQR))** | 1334 [1325, 1399] | 1346 [1299, 1434] | 1376 [1337, 1412] |
| **Global T2, ms (median (IQR))** | 51 [49, 53] | 52 [48, 54] | 49 [47, 53] |
| **T2 max, ms (median (IQR))** | 57 [55, 62] | 64 [62, 68] | 60 [57, 67] |
| **Global ECV, % (median (IQR))** | 25 [24, 28] | 26 [25, 28] | 25 [23, 27] |
| **ECV max, % (median (IQR))** | 30 [30, 34] | 31 [29, 35] | 30 [26, 34] |
| **Late gadolinium enhancement, n (%)** | 1 ( 20) | 4 (40) | 4 (40) |
| **Sub-endocardial LGE, n (%)** | 0 ( 0) | 2 (20) | 0 (0) |
| **Midwall LGE, n (%)** | 1 ( 20) | 3 (30) | 4 (40) |
| **RV insertion point LGE, n (%)** | 1 ( 20) | 2 (20) | 1 (10) |
| **Oxygen requirement, n (%)** | 1 ( 20) | 6 (60) | 8 (80) |
| **Myocarditis (MRI) (%)** | 1 ( 20) | 4 (40) | 3 (33) |
| **Myocarditis (PET) (%)** | 1 ( 20) | 5 (56) | 1 (11) |
| **Pulmonary inflammation, % of lung (median(IQR))** | 1 [1, 3] | 13 [4, 17] | 35 [29, 41] |

**Supplemental Table 9:** Imaging parameters stratified by pulmonary consolidation

**FIGURES**
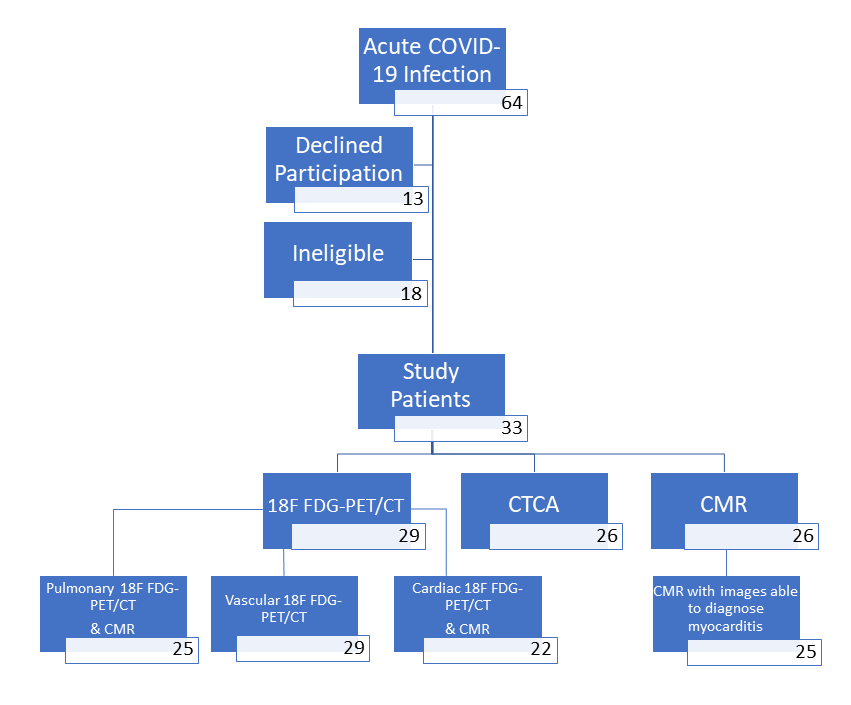

**Supplementary Figure 1**

Recruitment and scanning by multi-modality imaging. Of the 26 patients who had CMR, 1 had non-diagnostic T2 imaging so could not be used for assessment of myocarditis by the specific 2018 Lake Louis criteria. Of the 29 patients undergoing 18F-FDG PET/CT, all could be analysed for vascular inflammation. Of the remaing patients, 25 also had a CMR for comparison. Two patients were not adequately fasted for 18F-FDG myocardial analysis, and 1 CMR was non diagnostic, leaving 22 patients for comparison of cardiac pathology.

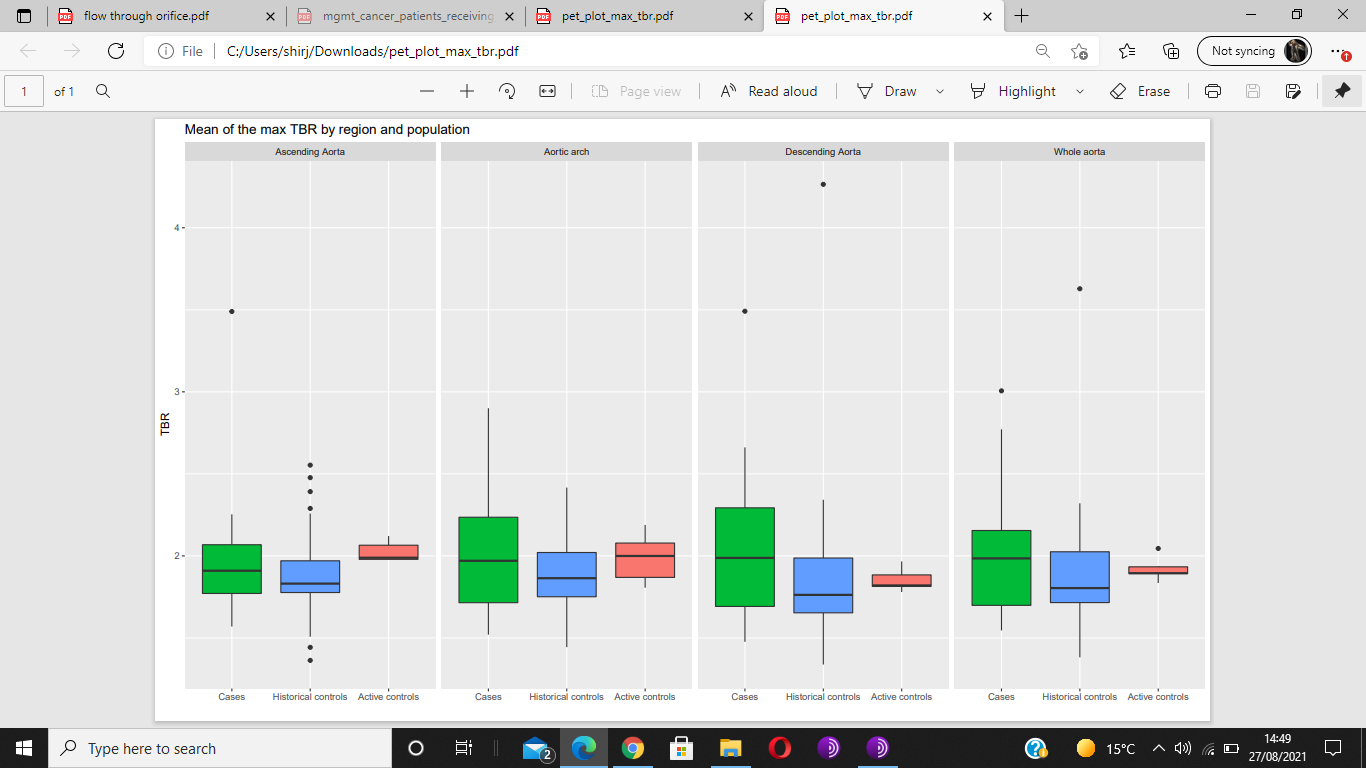

**Supplementary Figure 2**

Arterial inflammation in different regions of the aorta compared to active and historical controls.

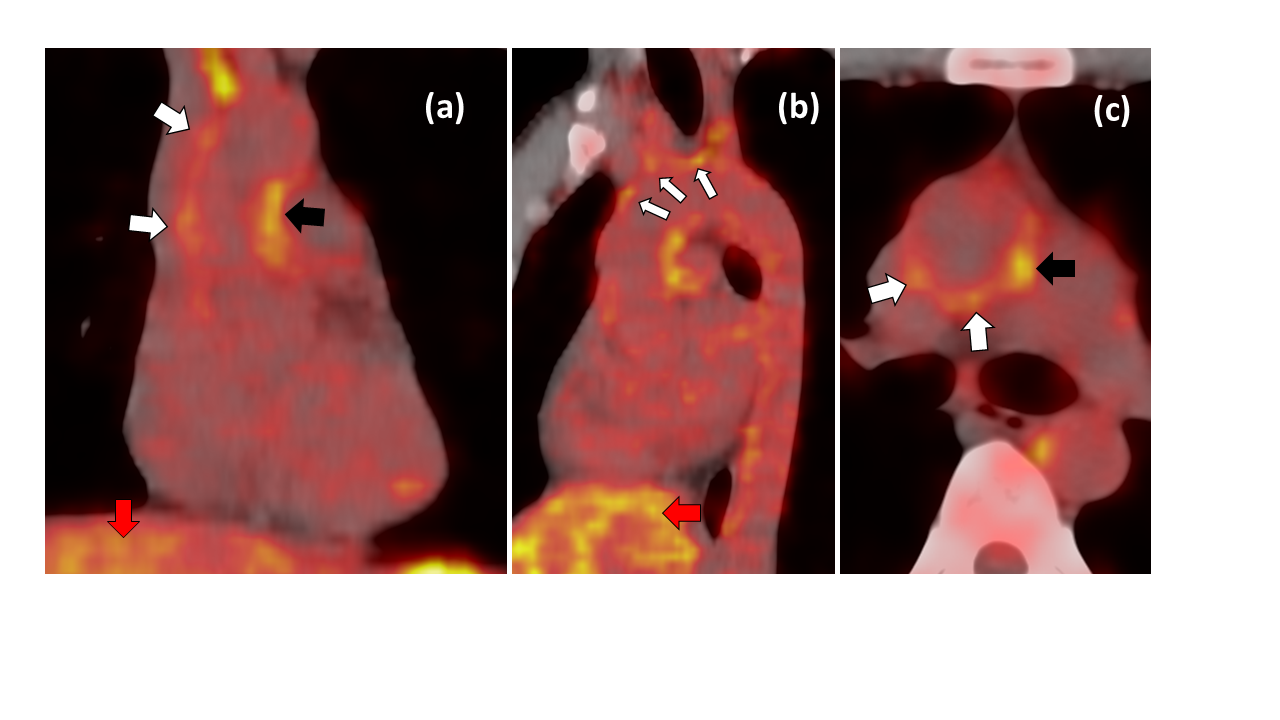

**Supplementary Figure 3**

18F-FDG PET uptake (white arrows) in the ascending aorta (a & b) and in the liver (red arrow). Liver uptake visually higher than the aortic uptake and by consensus this was graded at 1 using the American Society of Nuclear Cardiologists visual grading criteria**.** Black arrows may indicate possible brown fat uptake and was not considered diagnostic.
